# A Method to improve the Reliability of Saliency Scores applied to Graph Neural Network Models in Patient Populations

**DOI:** 10.1101/2022.04.06.22273515

**Authors:** Juan G. Diaz Ochoa, Faizan E Mustafa

## Abstract

Graph Neural Networks (GNN), a novel method to recognize features in heterogeneous information structures, have been recently used to model patients with similar diagnoses, extract relevant features and in this way predict for instance medical procedures and therapies. For applications in a medical field is relevant to leverage the interpretability of GNNs and evaluate which model inputs are involved in the computation of the model outputs, which is a useful information to analyze correlations between diagnoses and therapies from large datasets. We present in this work a method to sample the saliency scores of GNNs models computed with three different methods, gradient, integrated gradients, and DeepLIFT. The final sample of scores informs the customers if they are reliable if and only if all of them are convergent. This method will be relevant to inform customers which is the degree of confidence and interpretability of the computed predictions obtained with GNNs models.

## 1 Introduction

The widespread introduction of deep learning methods as an efficient method for performing inductive model induction has meant the fast development and adoption of black box models in their own nature, which has led to a considerable number of tools to improve the interpretability of this modelling method. A substantial portion of attention is focused on the saliency concept for deep learning, which consists of finding unique features in the context of the results delivered by the models [1]. While the concept of saliency has been applied in medicine to improve the interpretability of deep learning methods in radiology [2], this same concept can also be used for the interpretation of deep-learning methods in other medical fields. Here we applied saliency methods not only to increase the interpretability of graph neural networks (GNNs) used to model patient populations in order to predict medical procedures from existing ICDs and patient information [3], but also as a method to establish useful correlations between the ICDs and the procedures, encoded as therapy keys (TKs). Although many saliency methods have been standardized for AI interpretability and transparence, rigorous investigation of the accuracy and reliability of these strategies is necessary before they are integrated into the clinical setting [2]. In general, saliency methods could be unreliable [4], delivering false interpretations as, for instance, false correlations between ICDs and TKs.

This type of assessment is also helpful in assessing the uncertainty associated with certain predictions when salience methods are used, so that low uncertainty is achieved when all salience values calculated with the different methods converge to similar or equal values. This end result is relevant because we want to assess when a particular prediction and correlation is uncertain, i.e. is a method that is able to inform the customer when the assignment of ICDs to TKs is uncertain, and is an alternative to other methods aimed at improving the transparency of deep learning applications, for example medical recommendation systems. [5][6]. In addition, these results are helpful in evaluating the plausibility that the predicted TKs are really mapped to the input characteristics (in this case, ICDs). We are implementing our analysis in GNN models in synthetic patient populations with chronic disease, as Diaz Ochoa & Mustafa have reported [3].

## 2 Methods

We evaluate the predictions of TKs obtained with GNNs for a network derived from a synthetic patient database, with each patient assigned a series of ICDs describing their behavior, resulting in a patient graph structure based on basic patient information such as age and gender, as well as the diagnoses and trained GNNs models, to guide the patient’s therapies (see figure 1). The implementation as well as results are reported by Diaz Ochoa & Mustafa [3]. Despite this implementation demonstrated that the accuracy of GNNs is relatively high, in this implementation the correct relation between inputs (ICDs) and outputs (TKs) remains challenging.

**Fig 1.**
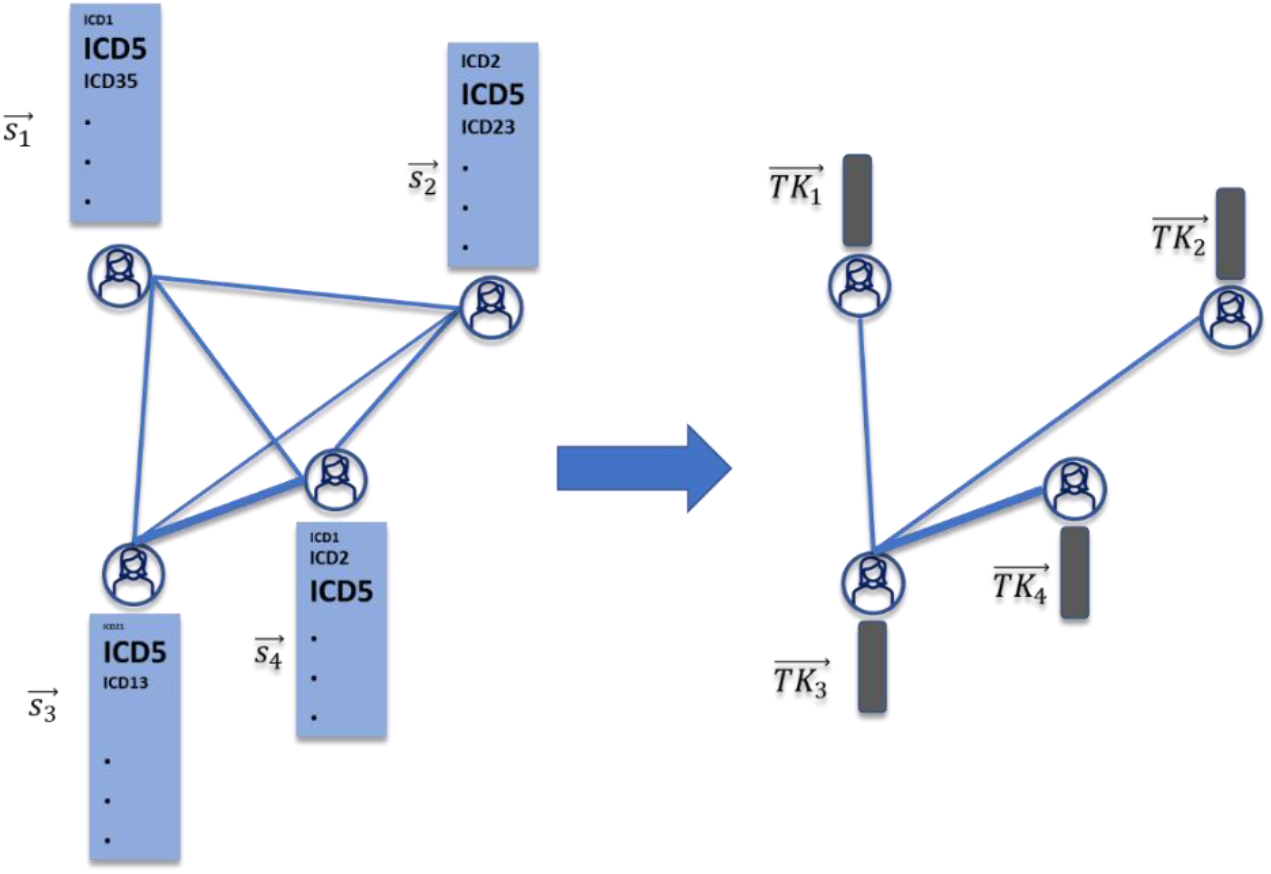
Intuitive interpretation of the meaning of the saliency score for a graph. The size of the elements in the ICD vector 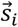 represent the saliency (importance) of the ICD for the prediction of the TKs,

Since the outputs are lists of TKs assigned to each patient, in this implementation was possible to identify a full network as well as seven characteristic clusters of patients with both similar TKs and ICDs, after predicting these values using the trained GNN model on test data [3]. The score of the input ICDs involved in the prediction of the label, in this case each TK, defined as 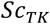, is in principle a non-linear function that can be approximated to a linear function, in this case the ICD vectors, such that 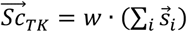 (intuitive interpretation in figure 1). This computation can be performed for the full graph, as well as for each one of the identified clusters *n* = {0,1,2,3,4,5,6}.

In a first instance the relation between input features and TKs has been approximated by a Taylor approximation with gradient methods.^1^ [7]. The score *w* can then be computed by back propagation, i.e., the gradient of the model output (difference of number of times a single label is observed) respect to the input (different single combinations of ICDs) 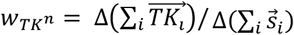 (from equation 3), which depend on the vector 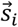 for each node (patient) *i* (see Fig. 1), where Δ is the difference between the number of predicted TKs and the difference of the number of ICDs in the graph. This difference is computed considering the frequency of ICDs in the sample. In this way we obtain an information about which ICDs are more important for the prediction of TKs. This score is computed either for the whole label-network as well as for each one of the identified sub-networks.

Furthermore, since sensitivity and implementation invariance are non-preserved in conventional gradient methods, we also implemented integrated gradient methods^2^ defined as the path integral of the gradients along the straight-line path from the baseline x′ to the input x [8].

Despite these methods are helping to better understand what a deep-learning method is “currently seeing”, several works have demonstrated that they are not reliable enough. For this reason, the DeepLIFT^3^ method has been obtained to leverage saliency analysis, which is a method that compares the activation of each neuron to its ‘reference activation’ and assigns contribution scores according to the difference [9]. This method has proven to be effective in cases when a neuron in the Neural Network, defined for instance by a ReLu activation function, carries information, despite it is not firing, such that the gradient in this case is equal zero.

Since we are dealing with three different methods delivering different scores, we approximated the scores into a single ranking of five levels (ranking from a low, medium and high saliency), such that the score computed with each method is grouped into 5 bins based on equal sized quartiles, i.e., we mapped the scores 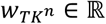, into a discrete scale 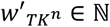 ranging from low 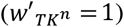 to high 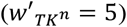 saliency. After applying this transformation for the scores obtained with all the three different methods, we can then sample these scores to perform the final analysis.

## 3 Results

The computed samples of 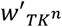 were estimated for the full network as well as for each one of the identified clusters. We have discovered that 27.2% of the saliency scores are convergent, considering that in the database we have 250 different ICDs. Furthermore, a large fraction of the predicted 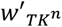 per ICD has a large skewness, i.e., in some cases one of the methods is delivering high divergent values respect the other two methods.

In the analysis of the full graph, we also obtained that 20% of the median was related to a large saliency value (importance) (median value equal to or greater than 3) and mainly the convergent salience values for “less important” ICDs were obtained (see Table 1). The size of the saliency or importance is however not related to the ICD’s frequency, i.e., how often an ICD is used (reflecting the number of times that a morbidity and comorbidity is coded). Thus, the way how the GNN model recognizes features in the ICD distribution of the patient population is not necessarily related to the ICD frequency, but it can be generated by the interdependence of features on each other.

**Table 1.** Accuracy of the saliency scores for each one of the obtained TK sub-graphs based on the GNN predicitons.

| $n$ | Percentage converging Saliency Scores (%) | High Scores: Relative % convergence – median $\geq 3$ | Low Scores: Relative % convergence – median $< 3$ |
| --- | --- | --- | --- |
| Full | 27.20 | 20.00 | 28.00 |
| 0 | 44.00 | 0.00 | 45.00 |
| 1 | 40.80 | 40.00 | 41.00 |
| 2 | 28.00 | 0.00 | 31.00 |
| 3 | 56.00 | 0.00 | 60.00 |
| 4 | 36.00 | 14.00 | 37.00 |
| 5 | 52.40 | 17.00 | 53.00 |
| 6 | 44.80 | 14.00 | 46.00 |

Although the result in Figure 2 is a good proxy to understand how the saliency scores are working in the graph, this calculation is less informative, in part because the reliability is relatively low and because there is a strong salience divergence for each of the ICDs. Instead, a salience analysis of the identified clusters of patients with similar TKs could be much more informative and accurate, using the results of the sub-graph analysis as reported in Diaz Ochoa & Mustafa [3].

**Fig 2.**
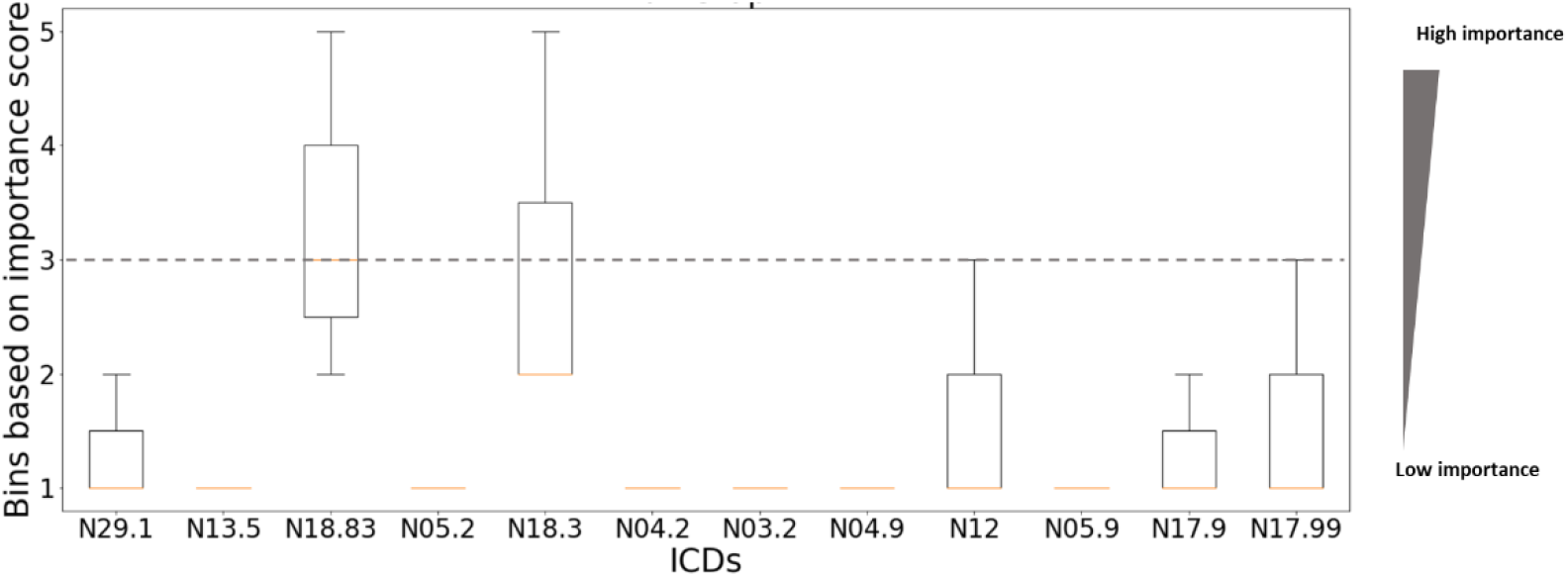
Boxplot of the bin-sampled saliency scores (y axis) as a function of ICDs (a representative fraction from 250 ICDs) for the full ICD-patient graph, as reported in [3]

In the next part we perform a similar saliency analysis for each one of the computed subclusters *n*. Interestingly, the convergence of scores in the subgraphs is better than for the full graph, with 56% of convergent scores for cluster number 3, i.e., as expected, it is much reliable to analyze the relationship between the model outputs and inputs for the subgraphs than for the full graph (see Fig. 3 as well as Table 1). Also observe that the saliency scores computed for the full graph might be different for the subgraphs (see for instance N18.83 in the full graph and in the subgraph 6). In general, with all three methods, we get a mean convergence of about 43%. This result is relevant because it implies that we are getting greater precision **and** interpretability for subgraph analysis in relation to full graph analysis. However, the reliability of the saliency is still too low, not only with respect to the whole ICD, but with respect to the total number of High and Low scores. In particular, due that the sample for high scores is smaller, we have obtained in this case for some subgraphs no convergence.

**Fig. 3.**
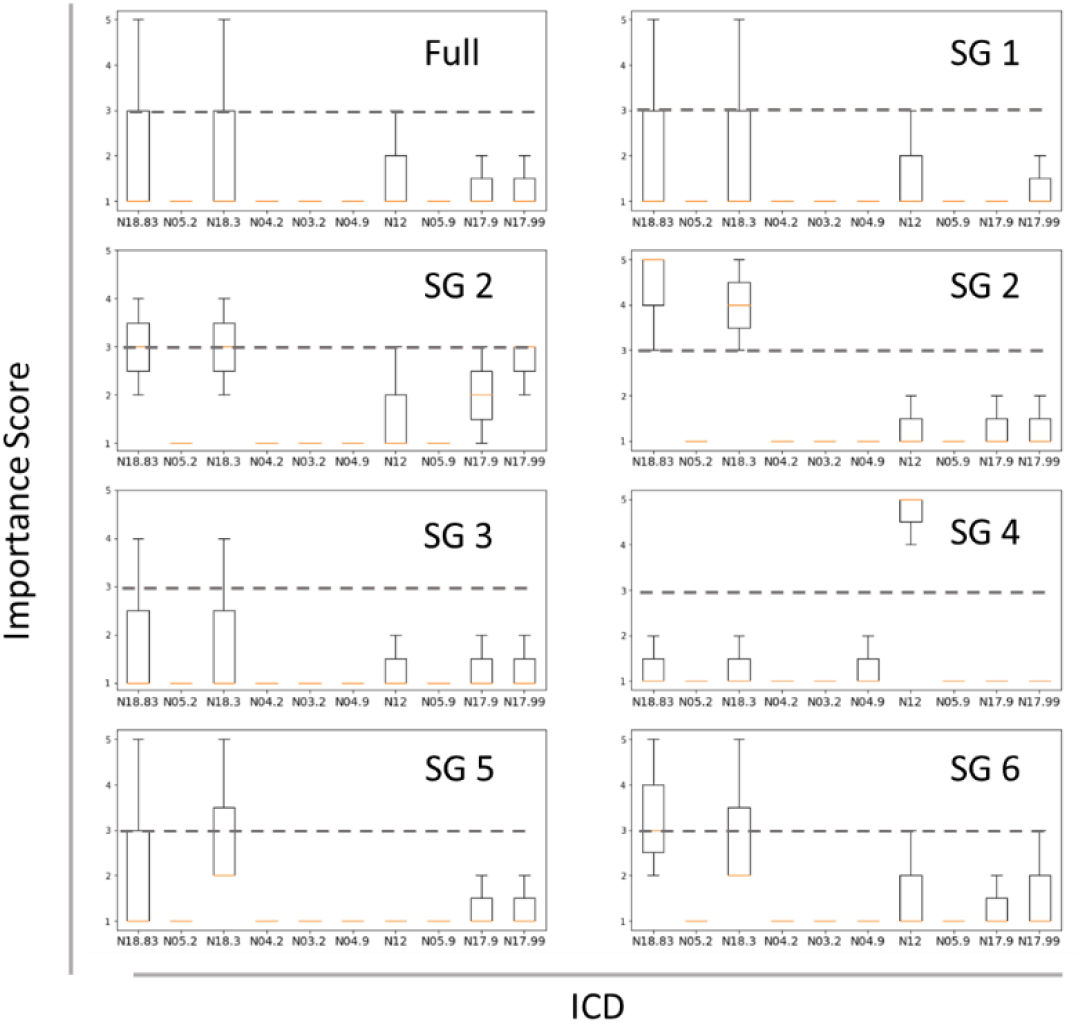
Boxplot distribution of the computed saliency scores (y axis) after clustering similar TKs as a function of the ICDs (x axis, only a selected number) for different identified sub-graphs SG.

These results have two implications:

- **First**, the saliency analysis is much better in subgraphs identifying specific TK clusters, i.e., the saliency method has a good functionality when there is a guarantee that the GNNs has a given specificity.
- **Second**, the obtained results imply that either all the three saliency methods are unreliable, impairing the estimation of the correlations between ICDs and procedures, or that this estimation is essentially difficult to be performed due to the irregular and variable assignment of medical procedures (epistemic uncertainty).

In this case this result can be used in two ways: one, to establish the saliency score and in this way estimate which ICD combination could be involved in the estimation of a procedure, and second use the degree of uncertainty in the scores (skewness and distribution) to inform the customer both if the prediction and ICD-OPS correlation computed with the model is robust or uncertain and how significative could be this uncertainty when the computed TKs are related to the inputs. Thus, this method is an alternative approach to inform customers about the effect of epistemic uncertainty in the original graph-database on the final model outputs.

## 4 Conclusions

In this work, we developed a method of analyzing different salience methods to evaluate their reliability when applied to GNNs that encode similar ICDs in a patient population with a chronic disease to predict their TKs. The salience method is important both to improve the interpretability of the implemented GNN and secondly to calculate the exact correlation between TKs and ICDs.

First, the developed method is a proof of concept to take advantage of the reliability of the calculated saliency scores and simultaneously to inform the customers of the model about potential epistemic uncertainties and uncertain interpretability of point predictions, in this case the assignment of ICDs to TKs.

Second, the calculated results show that using salience values in partial graphs is more effective than in full graphs. Since the combination of morbidities and comorbidities is better represented in the subgraphs and subclusters than in the complete graph, the calculation of the salience values can be more effective for these subgraphs. These results will guide the application of GNNs in the design of, for example, recommender systems in medicine, taking into account not only technical but also relevant ethical aspects that determine how model results, and corresponding inaccuracies and uncertainties, should be communicated to the customer [10].

## Data Availability

All data produced in the present study are available upon reasonable request to the authors

## Acknowledgments

We would like to thank M. Grundman and T. Schimper for successful exchange and information, and F. Weil for his constant support in the realization of this project.

## Data availability and ethical issues

The data used in this study is fully synthetic. All data produced in the present study are available upon reasonable request to the authors

## Footnotes

1 https://captum.ai/api/saliency.html

2 https://captum.ai/api/integrated_gradients.html

3 https://captum.ai/api/input_x_gradient.html

## Notes

### Competing Interest Statement

The authors have declared no competing interest.

### Funding Statement

This study did not receive any funding

